# Self-blame-selective social conceptual overgeneralization and vulnerability to depression

**DOI:** 10.1101/2025.05.06.25327075

**Authors:** Diede Fennema, Andrew J. Lawrence, Catherine A. Spilling, Jorge Moll, Matthew A. Lambon Ralph, Roland Zahn

## Abstract

**Background:** Beck observed overgeneralized thinking as a key vulnerability factor for excessive self-blame/-criticism in major depressive disorder (MDD). Whilst the contribution of reduced access to specific autobiographical memory episodes has often been considered, the role of more abstract semantic memory systems remains elusive. Here, we investigated the individual ability to differentiate between the meaning of abstract social concepts when interpreting behavior (e.g. “critical” vs. “faultfinding”) and its contribution to vulnerability to self-blaming biases and MDD using a previously developed cognitive task.

**Methods:** Cognitive testing in 96 participants (n=60 medication-free remitted MDD and n=36 control) and fMRI scanning in 75 participants employed self- and other-blame-evoking conditions. *A priori* right anterior temporal lobe (ATL) seed and bilateral anterior subgenual cingulate and dorsolateral prefrontal cortex regions-of-interest were defined.

**Results:** As expected, the MDD group exhibited greater self-blame-selective conceptual overgeneralization and stronger interdependency of conceptual overgeneralization with negative emotional valence relative to the control group. Individuals with this depressogenic interdependency showed higher self-blame-selective right anterior subgenual cingulate and primary motor cortex activations across groups, potentially corresponding to stronger self-blame and self-agency attributions, respectively, when conceptually overgeneralizing the interpretation of their negative actions. Individuals with higher self-blame-selective conceptual overgeneralization displayed lower right ATL activation for self-vs. other-blame, suggesting reduced access to differentiated conceptual representations.

**Conclusions:** Future studies are needed to confirm the hypothesis that self-blame-selective conceptual overgeneralization characterizes a distinct neurocognitive subtype of primary MDD vulnerability which may be modulated by depressive state and thus serves as a personalized treatment target.

## Introduction

The revised learned helplessness model posits that when an individual attributes the cause of failure to their unchangeable global character faults, this renders them vulnerable to feelings of self-worthlessness and major depressive disorder (MDD) (1, 2). Self-criticism-related overgeneralization is a prominent feature of MDD, and is targeted by cognitive psychotherapy (3, 4). Unveiling the neurocognitive mechanisms underpinning individual vulnerability to overgeneralized thinking is therefore key to a comprehensive understanding of MDD.

Overgeneralized autobiographical memory biases have been established as transdiagnostic mechanisms relevant to affective disorders such as MDD and trauma-related disorders (5). Whilst the contribution of reduced access to specific autobiographical memory episodes has often been considered in these studies, the role of more abstract semantic memory systems is unclear. Abstract social concepts such as “rude” and “vain” are used to describe so-called “personality” traits people attribute to themselves or others and are intimately linked with one’s self-worth and its relationship with depression (6). Social conceptual differentiation is thought to allow for a narrower interpretation of situations (e.g. “clumsy”), thereby protecting from overgeneralizing to other aspects (“I am a total failure”).

Using a task probing differentiated conceptual knowledge, Green *et al.* (7) provided preliminary evidence for the relevance of conceptual overgeneralization to MDD vulnerability in people with remitted MDD (rMDD). They found a stronger relationship between overgeneralizing social concepts and negative emotional valence of these concepts, for rMDD participants compared to the control group. In other words, the failure to discern nuances in the meaning of social concepts was accompanied by perceiving the social actions as more unpleasant in the rMDD group, which suggests that conceptual overgeneralization plays a role in the experience of emotions.

Previous work has identified the anterior temporal lobes (ATLs) as core to representing differentiated conceptual knowledge (8–12). The right superior ATL was hypothesized to represent conceptual aspects of social behavior, as evidenced by functional MRI (fMRI) studies showing a stronger activation when contrasting social vs. non-social concepts as well as associations with conceptual detail and semantic relatedness (13). More pronounced impairments for social vs. non-social concepts were observed in patients with right superior anterior temporal dysfunction due to frontotemporal lobar degeneration (14, 15) and when applying repetitive transcranial magnetic stimulation interfering with right superior ATL function (16). However, when considering frontotemporal lobar degeneration with advanced bilateral ATL volume reductions, both abstract non-social and social concepts were found to be impaired (17).

The right superior ATL has been implicated in self-blaming emotions through its coupling with septal-subgenual regions, which is likely to facilitate integration of conceptual information about the social meaning of a situation (as represented in the right superior ATL) with agency-context-related information (as represented in the subgenual region) (18–20). Green *et al.* (7) showed reduced functional right superior ATL-subgenual coupling in a rMDD group, suggesting that impaired conceptual-emotional integration could lead to a depressogenic attributional style (1, 2, 21).

Here, we aimed to provide more definitive evidence in a larger sample and expand Green *et al.*’s (7) findings. Firstly, we predicted that the rMDD group would show self-blame-selective conceptual overgeneralization, i.e. differentiate social concepts less when appraising their own compared to others’ negative social actions. We expected that, as previously, the rMDD group would show stronger interdependence of self-blame-selective conceptual overgeneralization and associated negative emotional intensity, which could explain the observation that people with MDD feel more intense depressive affect when they overgeneralize self-critical content (21).

Secondly, we hypothesized that self-blame-selective overgeneralization would be associated with self-blaming biases. Green *et al.* (7) showed that individuals with higher interdependence of conceptual overgeneralization and negative emotional biases exhibited stronger self-hate on the Interpersonal Guilt Questionnaire (22). Here, we investigated a more selective form of self-blaming bias validated in our more recent work in MDD, i.e. reduced proneness to externalized anger leading to a relative increase in self-vs. other-blame-related emotions (23, 24). Interestingly, irritability (i.e. externalized anger proneness) was observed in a subgroup of MDD (25–27), potentially as an intact self-defense mechanism, which we hypothesized to be associated with the absence of self-blame-selective conceptual overgeneralization.

Finally, based on Green *et al* (7), we hypothesized that stronger conceptual-emotional interdependence would be associated with reduced right superior ATL connectivity with frontal-subcortical circuits, specifically, subgenual anterior cingulate and dorsolateral prefrontal cortex (DLPFC). The subgenual region has been reproducibly associated with individual differences in proneness to self-blaming feelings (23), while DLPFC dysfunction is thought to contribute to MDD symptoms (28, 29), potentially due to its role in reappraisal of emotions (30) by representing sequential action/goal contexts (31).

## Methods and Materials

The cognitive task data first reported here, were collected as part of a previously published study which examined whether self-blame-selective alterations in anterior temporal fMRI connectivity predicted subsequent recurrence of depression (19). Functional MRI results regarding blood-oxygen level-dependent (BOLD) subgenual cingulate cortex activation and proneness to self-blaming emotions, as well as internal reliability of self-blame-related fMRI measures have been described elsewhere (32, 33).

### Participants

Participants in the rMDD group fulfilled criteria for MDD according to Diagnostic and Statistical Manual of Mental Disorders (34), without any current Axis-I disorders, and had been in remission for at least six months. The control group had no current or past Axis-I diagnoses and no first-degree family history of mood disorders. Both groups were psychotropic medication-free. Ethical approval was obtained from the South Manchester National Health Service Research Ethics Committee (ref: 07/H1003/194). All participants provided informed consent and received compensation. For full details, see Supplementary Methods.

In total, 160 participants (108 rMDD; 52 control) were included. Of those, 96 participants (60 rMDD; 36 control) had complete task data, and 75 participants (45 rMDD; 30 control) also had complete fMRI data with good coverage of ventral frontal, frontopolar and ATLs, and acceptable range of head motion (translation<6 mm, or rotation<4 degrees). There were no significant group differences in education, age or sex, but the rMDD group had higher residual depressive symptoms and lower psychosocial functioning than the control group (Table 1).

**Table 1.** Demographic and basic clinical characteristics.

|  | Remitted MDD | Control group | Total | Comparison MDD vs controls |
| --- | --- | --- | --- | --- |
|  | n = 60 | n = 36 | n = 96 |  |
| Sex | 40 female / 20 male | 22 female / 14 male | 62 female / 34 male | $\chi^2(1,96) = .30, p = .58$ |
| Age in years | 36.9 ± 13.3; 18 - 63 | 34.5 ± 14.3; 20 - 64 | 36.0 ± 13.7; 18 - 64 | $t(94) = .82, p = .21$ |
| Years of education | 16.9 ± 2.2; 12 - 22 | 17.5 ± 2.5; 13 - 25 | 17.1 ± 2.3; 12 - 25 | $t(94) = -1.23, p = .11$ |
| BDI score | 3.9 ± 3.9; 0 - 15 | 1.0 ± 1.7; 0 - 6 | 2.8 ± 3.5; 0 - 15 | Mann-Whitney $U(96) = 521.0, p < .001^*$ |
| MADRS | 1.0 ± 1.5; 0 - 4 | 0.7 ± 1.3; 0 - 4 | 0.9 ± 1.4; 0 - 4 | $t(94) = 1.07, p = .29$ |
| GAF | 85.4 ± 6.3; 70 - 90 | 89.0 ± 2.8; 80 - 91 | 86.8 ± 5.5; 70 - 91 | Mann-Whitney $U(96) = 1436.0, p = .001^*$ |
| Number of lifetime MDEs | 4.2 ± 7.1; 1 - 53 | | | |

\* significant at $p < .05$ , two-tailed. Means, standard deviations and range are reported ( $M \pm SD$ ; *minimum – maximum*). MDD = major depressive disorder; BDI = Beck Depression Inventory; MADRS = Montgomery-Åsberg Depression Rating Scale; GAF = Global Assessment of Functioning Scale; MDEs = Major Depressive Episodes.

### Clinical measures

Residual depressive symptoms and psychosocial functioning were measured using the Beck Depression Inventory (BDI) (35), Montgomery-Åsberg Depression Rating Scale (MADRS) (36) and Global Assessment of Functioning Scale (GAF) (34). The number of previous major depressive episodes was used an indicator of potential “scarring” effects of repeated depressive episodes, and anger towards others on the Value-related Moral Sentiment Task (VMST) as a measure of self-blaming bias (see Supplementary Methods).

### Conceptual social knowledge differentiation (CSKD) task

During the CSKD task (described in full in Green *et al.* (7)), participants were presented with short written statements describing hypothetical social behaviors. Participants completed two versions of the task, one comprising negative social behaviors and one comprising positive social behaviors. The statements differed in their agency with either the participant (self-agency; 30 items) or their best friend (other-agency; 30 items) acting negatively or positively towards the other. For example, participants were asked to imagine “[participant’s name] ignoring [best friend’s name] at a social event” (self-agency) and “[best friend’s name] ignoring [participant’s name] at a social event” (other-agency). Participants were instructed to select the social concept that best described the hypothetical social behavior, from a list of 19 concepts, e.g. “boastful” or “jealous”, and to only choose more than one concept if it described the behavior equally well. Therefore, choosing more than one concept indicated conceptual overgeneralization (7). Participants were also asked to rate the valence of the social behavior ranging from −4=extremely unpleasant and 4=extremely pleasant. Based on Green *et al.* (7), we focused on the negative CSKD version for our primary analyses (see Supplementary Materials for the positive social behavior version).

### Behavioral data analysis

The task was designed to investigate differences in conceptual differentiation depending on agency-role (self vs. other). As described in Green *et al.* (7), CONCEPT scores were calculated as the number of concepts chosen in the other-agency condition subtracted from the number of concepts chosen in the self-agency condition (here, we used means rather than sum scores to be more robust against missing trials in future studies), as a measure of relative indistinctiveness of concepts in one condition relative to the other. VALENCE scores were computed as the differences in mean pleasantness ratings between self-agency and other-agency conditions, i.e. emotional valence. The CONCEPT x VALENCE interaction was calculated as the product of CONCEPT and VALENCE scores.

All data analyses were carried out using IBM SPSS Statistics 29 (IBM Corp., Armonk, NY). Group differences in CSKD scores, correlations with clinical measures and a multivariate analysis of variance investigating the relationship between CSKD scores and risk of recurrence were performed (see Supplementary Methods). Results were considered significant at *p*=.05 (two-tailed).

### Functional MRI methods

#### fMRI acquisition and paradigm

As previously described (18), we used an fMRI protocol designed to improve sensitivity within ventral brain regions (see Supplementary Methods). Participants were shown short written statements describing hypothetical social behaviors, in which either the participant (self-agency; number of trials=90) or their best friend (other-agency; number of trials=90) acted contrary to socio-moral values described by a social concept (e.g. impatient, dishonest). Stimuli were presented for five seconds in three runs in pseudorandom order; runs were counterbalanced across participants and were interspersed with a baseline visual fixation pattern condition (number of trials=90), followed by a jittered inter-trial interval with a mean duration of four seconds. In the scanner, participants were asked to decide whether the described behavior would feel mildly or very unpleasant. For full details, see Supplementary Methods.

#### fMRI analysis

At the individual level, we modeled standard BOLD effects for self- and other-agency conditions, following a similar approach as previously described (19, 32, 33) (for full details, see Supplementary Methods). We modeled connectivity using psychophysiological interaction (PPI) analysis, which required the extraction of the signal time course within the right superior ATL seed region (same as Green *et al.* (7); 6mm sphere, centered on Montreal Neurological Institute [MNI] coordinates: x=58, y=0, z=12). Interaction terms were calculated with the main effects of self-agency vs. fixation and other-agency vs. fixation.

At the second level, one-sample *t*-tests were performed comparing BOLD and PPI contrast maps for self-vs. other-blaming emotions, while modelling CONCEPT, VALENCE and CONCEPT x VALENCE scores as covariates. All analyses were inclusively masked using grey matter masks derived from spatially normalized T1-weighted images (18).

Statistical inference was made at the voxel-level using a threshold of *p*=.001 (uncorrected), with subsequent Family-Wise Error (FWE) correction at *p*=.05 across the whole-brain, and with small-volume correction across our *a priori* defined regions-of-interest (ROI, see below). Using the MarsBaR toolbox (37), we extracted cluster averages from regions emerging from the whole-brain analysis and ROI averages for individual participants for the subtraction-based difference contrast (self-minus other-agency), for exploratory analysis in IBM SPSS Statistics 29.

#### fMRI ROIs

Our *priori* ROIs were based on previous independent studies showing relevance for social conceptual-emotional integration, i.e. part of the right ATL, bilateral DLPFC, and bilateral subgenual anterior cingulate (see Supplementary Methods).

## Results

### CSKD task results

Compared with the control, the rMDD group showed self-blame-selective conceptual overgeneralization, i.e. selecting more concepts when appraising negative actions in the self-vs. other-agency condition (CONCEPT scores: rMDD, M=0.20, SD=0.43; control, M=0.02, SD=0.20; *t*[89.2]=2.81, *p*=.006; see Figure 1 for an example of an rMDD participant). We replicated Green *et al.* (7)’s finding of more negative CONCEPT x VALENCE scores in rMDD vs. control groups (rMDD, M=-0.15, SD=0.42; control, M=-0.02, SD=0.08; *t*[65.1]=-2.30, *p*=.03, reflecting more positive CONCEPT scores being associated with more negative VALENCE scores), reflecting that individuals with rMDD with self-blame-selective conceptual overgeneralization associated more negative valence with the self-agency condition. However, as before, there was no group difference in VALENCE scores (rMDD, M=-0.18, SD=0.48; control, M=-0.13, SD=0.42; *t*[94]=-0.54, *p*=.59). Consistent with Green *et al.* (7), the positive social behavior showed no group differences in CONCEPT, VALENCE or CONCEPT x VALENCE scores (see Supplementary Results).

**Figure 1.**
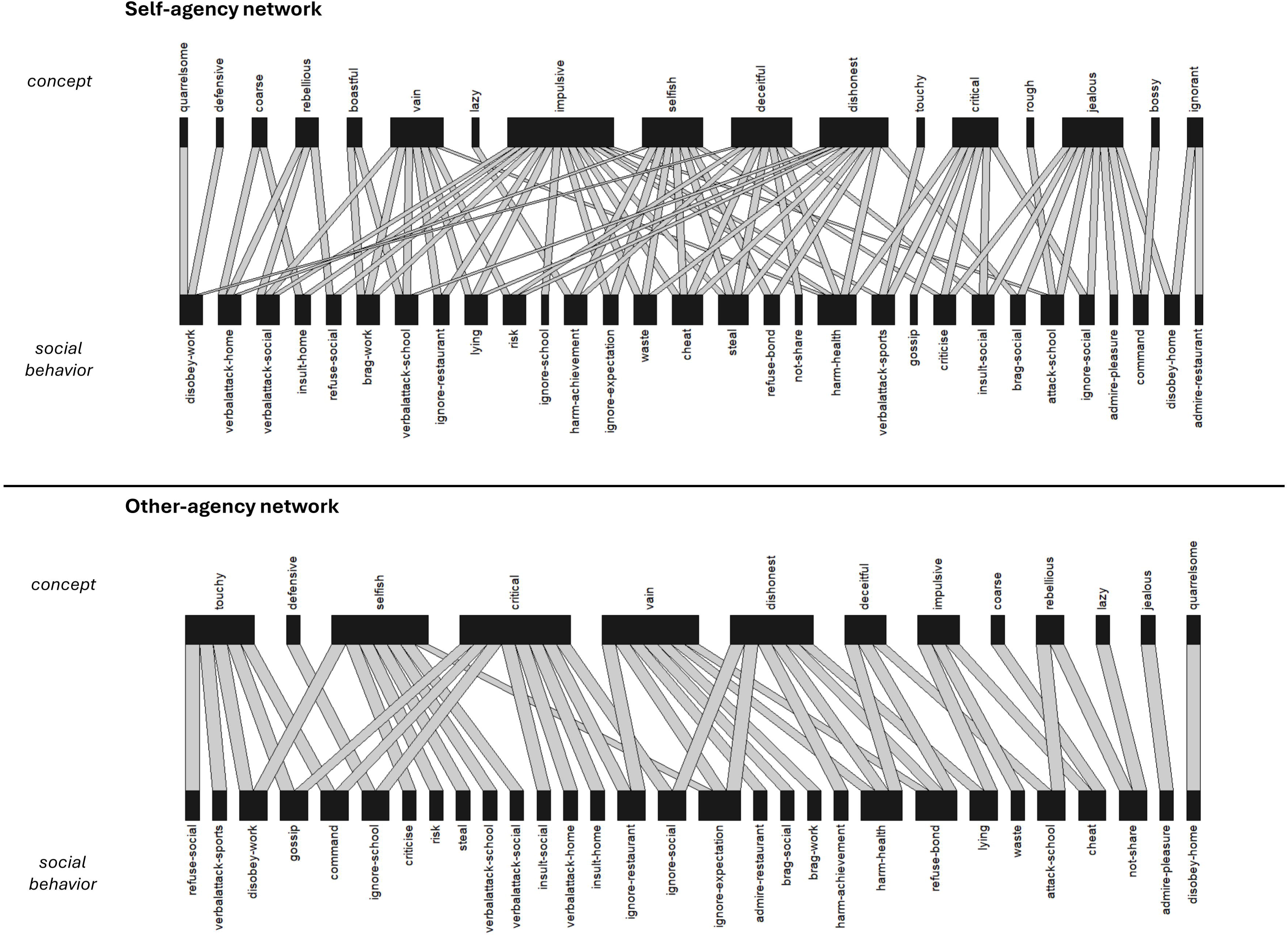
**Visualization of conceptual knowledge bipartite network of self- and other-agency conditions in a participant with remitted MDD.** We examined the relationship between concepts and social situations by treating it as a bipartite (two-mode) network, given a concept cannot connect to another concept except through the social situation. Degree centrality, which gives a value to the number of lines connecting the concepts to the various social situations, was computed using the R package “igraph” (68). As expected, the remitted MDD group showed higher degree centrality in the negative self-agency condition (M=4.18, SD=1.68) than in the other-agency condition (M=3.70, SD=1.31; *t*[59]=3.56, *p*<.001). In contrast, the control group displayed similar mean degree centralities in the self-agency (M=3.43, SD=1.11) and other-agency conditions (M=3.39, SD=1.23; *t*[35]=0.48, *p*=.63). The figure shows the connections between the 19 concepts and 30 social behaviors in the self-agency and other-agency condition by one participant with remitted MDD (using the R package “bipartite”) (69). This participant used more concepts as descriptive of their own (n=75) relative to their best friend’s behavior (n=46), indicating conceptual overgeneralization for self-relative to the other-agency condition. MDD = major depressive disorder.

### fMRI results

Contrary to our hypothesis, there was no self-blame-selective right superior ATL connectivity with our *a priori* ROIs or at the whole-brain level. However, our voxel-based BOLD analysis revealed a negative association between CONCEPT scores and the right ATL ROI, extending into the perirhinal cortex (BA20/36; Table 2; Figure 2), and there was a side-peak in the temporopolar part (BA38; Table S1). Using the combined whole-brain derived cluster averages of these two peaks, ATL BOLD activation showed a negative association with CONCEPT scores (*r*[75]=-.30, *p*=.01). In a supporting conjunction analysis, we found consistent bilateral BOLD effects across individuals in the bilateral superior ATL across the self- and other-agency conditions (Figure S1).

**Figure 2.**
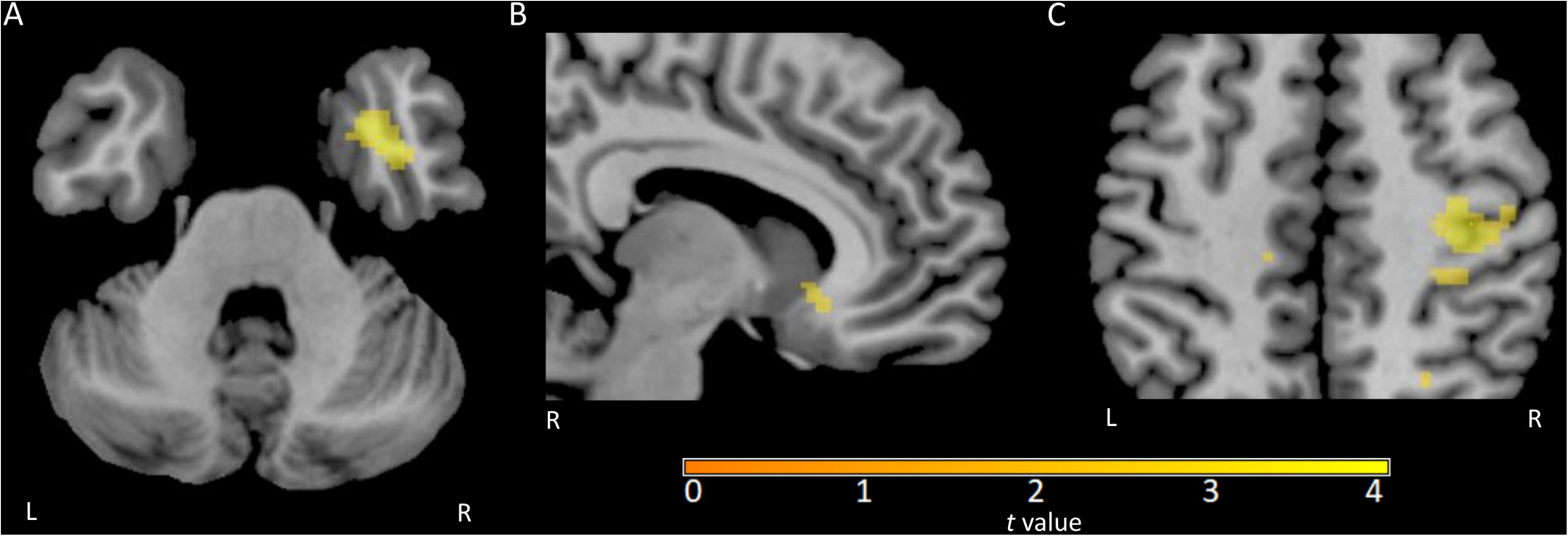
**Individual differences in CSKD task performance and BOLD effects** This figure shows cropped sections from n=75 participants through **(A)** anterior temporal lobe (ATL; BA20/36; MNI z-coordinate = −34), **(B)** anterior subgenual cingulate (BA24; MNI x-coordinate = 8) and **(C)** primary motor area (BA4/3; MNI z-coordinate = 50), displayed using MRIcron (70) at an uncorrected voxel-level threshold of *p* = .005, with a cluster-size threshold of 10. The color bar represents *t* values. The ATL **(A)** showed a negative association with CONCEPT scores (calculated as the difference in the mean number of concepts chosen for self-vs. other-agency), i.e. those with self-blame-selective conceptual overgeneralization showed lower right ATL BOLD activation. In contrast, the anterior subgenual cingulate and primary motor area showed a negative association with CONCEPT x VALENCE interaction scores (where VALENCE was calculated as the difference in mean valence ratings (on a bipolar scale) in the self-vs. other-agency condition, and CONCEPT x VALENCE was computed by multiplication of CONCEPT by VALENCE scores). This means that those displaying increased BOLD activation for self-vs. other-blaming emotions, exhibited more negative CONCEPT x VALENCE interaction scores. As negative CONCEPT x VALENCE scores characterized the MDD relative to the control group, one can deduce that depressogenic CONCEPT x VALENCE interaction scores were associated with higher anterior subgenual cingulate and primary motor area activations. These negative interaction scores arose because in the MDD group, negative VALENCE scores, reflecting self-negative biases, were more strongly associated with positive CONCEPT scores across individuals, reflecting self-blame-selective overgeneralization. CSKD = conceptual social knowledge differentiation; BOLD = blood-oxygen level-dependent; ATL = anterior temporal lobe; BA = Brodmann Area; MNI = Montreal Neurological Institute.

**Table 2.** Individual differences in CSKD task performance and BOLD effects.

|  |  |  |  | MNI peak coordinates |  |  |  |  |
| --- | --- | --- | --- | --- | --- | --- | --- | --- |
| Hemi-sphere | Region | Cluster size | Brodmann Area | x | y | z | t statistic | Voxel-based FWE-corrected p value |
| BOLD negative effect of CONCEPT scores: |  |  |  |  |  |  |  |  |
| right | Anterior temporal lobe | 39 | 20/36 | 40 | -2 | -34 | 4.58 | .006<sup>a</sup> |
| BOLD negative effect of CONCEPT x VALENCE scores: |  |  |  |  |  |  |  |  |
| right | Anterior subgenual cingulate | 14 | 24 | 8 | 22 | -8 | 3.34 | .026<sup>b</sup> |
| right | Primary motor area | 80 | 4/3 | 36 | -22 | 50 | 5.42 | .018<sup>c</sup> |

<sup>a</sup> Region surviving voxel-based FWE correction over *a priori* right anterior temporal lobe region-of-interest as described in the supplement of Zahn *et al.* (46).
<sup>b</sup> Region surviving voxel-based FWE correction over *a priori* bilateral anterior subgenual cingulate region-of-interest (6 mm sphere, MNI: x=+/-4, y=23, z=-6, Green *et al.* (18).
<sup>c</sup> Region surviving whole-brain voxel-based FWE correction of *p* = .05.
Based on n=75 participants. CONCEPT is calculated as the difference in mean number of concepts chosen in the self- minus the other-agency as an index of conceptual overgeneralization; VALENCE is calculated as the difference in mean pleasantness ratings in the self- minus the other-agency as an index of emotional valence bias; and CONCEPT x VALENCE is the multiplication of CONCEPT and VALENCE scores. BOLD = blood-oxygen level-dependent; FWE = Family-Wise Error; MNI = Montreal Neurological Institute; CSKD = Conceptual Social Knowledge Differentiation.

Moreover, there was a negative association between CONCEPT x VALENCE scores and the *a priori* bilateral anterior subgenual cingulate ROI (BA24; Table 2; Figure 2). Using whole-brain derived cluster averages, this association appears to be mostly driven by the right anterior subgenual cingulate (*r*[75]=-.31, *p*=.01). Whole-brain level BOLD analysis revealed a negative association between CONCEPT x VALENCE scores and the right primary motor area (BA3/4; Table 2; Figure 2), which was confirmed when extracting the cluster averages (*r*[75]=-.37, *p*<.001).

There was no BOLD effect for our other pre-registered dorsal prefrontal cortex ROI. We also did not find any BOLD activations associated with VALENCE scores, or a positive association with CONCEPT scores or CONCEPT x VALENCE scores.

### Supporting analyses

#### Association between CSKD and clinical measures

Within the rMDD group, we found associations between some of the CSKD and clinical measures (Supplementary Results; Table S2). However, these associations are likely to have been driven by outliers, as only one association was robust to excluding outliers: showing that patients with self-blame-selective conceptual overgeneralization had lower psychosocial functioning (CONCEPT score and GAF rating: *r*[56]=-.30, *p*=.03). There were no correlations between the CSKD measures and any of the other clinical variables (Table S2). A multivariate general linear model with the three CSKD measures as outcomes found no significant effect of recurrence risk group, i.e. an ordinal variable with three levels (developing a recurrent episode over 14 months of follow-up (n=34), with subthreshold symptoms (n=9), and remaining stable (n=15; *F*[6,106]=1.19, *p*=.32; Wilk’s lambda=0.88; see Supplementary Methods for more details)).

#### Characteristics of “self-blame-selective conceptual overgeneralizer”

About half of participants (48%, n=29) with rMDD were classified as “self-blame-selective conceptual overgeneralizer” (i.e. CONCEPT scores>0), while 30% (n=11) of the controls showed self-blame-selective conceptual overgeneralization. The “self-blame-selective conceptual overgeneralizers” and “non-self-blame-selective conceptual overgeneralizers” did not differ in years of education, age, sex, residual depressive symptoms (BDI, MADRS) or psychosocial function scores (GAF; Table S3). There also was no association between group and self-blame-selective conceptual overgeneralization classification (χ^2^[1,96]=2.93, *p*=.09). However, “self-blame-selective conceptual overgeneralizers” showed a self-blaming emotional bias by exhibiting reduced proneness to feeling anger towards others on the VMST (M=17.7, SD=12.9) compared to “non-self-blame-selective conceptual overgeneralizers” (M=25.5, SD=15.0; *t*[93]=-2.65, *p*=.01).

## Discussion

As predicted, the rMDD group exhibited greater self-blame-selective conceptual overgeneralization, indicating a proneness to interpreting self-blame-related actions in an overgeneralized manner, as proposed by the revised learned helplessness model (1). Furthermore, we replicated the finding that interdependence of self-blame-selective conceptual overgeneralization and emotional valence was stronger in the rMDD group compared with the control group. Thus, when conceptually overgeneralizing, people with rMDD felt more negatively about the described behavior than control participants.

Interestingly, self-blame-selective conceptual overgeneralization characterized about half of the participants with rMDD and about a third of the control participants. The observation that self-blame-selective conceptual overgeneralization was not exclusive to the MDD group, suggests that other factors play a role in the development of MDD. Previous work proposed that multiple mechanisms may interact to contribute to self-blaming biases, and that conceptual overgeneralization is a contributing, but not necessary, factor (18). Complementary cognitive systems are involved in the appraisal of social behaviors (38). For instance, retrieval of specific, emotionally relevant autobiographical episodes may serve to counteract overgeneralized thinking (39).

Previous work has shown that an imbalance of specific self- and other-blaming emotions, rather than a general increase in negative emotions, is associated with vulnerability to MDD (24, 40). Specifically, MDD demonstrated a relative reduction in negative emotions when blaming others. In keeping with this, we also found that self-blame-selective conceptual overgeneralizers showed reduced externalized anger, as an expression of self-blaming biases, relative to conceptual non-overgeneralizers. The ability to externalize anger towards others may represent an intact self-defense mechanism to protect one’s self-esteem (41), which could be weakened by self-blame-selective conceptual overgeneralization.

Even though our participants were in full remission, self-blame-selective conceptual overgeneralization was associated with lower psychosocial functioning scores, irrespective of residual depressive symptoms. This suggests that self-blame-selective conceptual overgeneralization could be clinically relevant, especially as other forms of overgeneralization have been shown to predict depressive symptoms (42, 43). Cognitive bias modification procedures, such as rumination-focused cognitive behavioral therapy or training patients to improve the specificity of their emotional recollection, have already been applied in the context of overgeneralized memory (44). Future research could focus on developing interventions targeting self-blame-selective conceptual overgeneralization.

We were unable to replicate Green *et al.*’s (7) fMRI findings which were based on functional/effective connectivity measures, likely due to the complex nature of these measures and poor reliability (33, 45). BOLD effect analyses, however, found more reliable than connectivity measures (33), revealed that individuals with lower self-blame-selective activation of the right ATL exhibited higher self-blame-selective conceptual overgeneralization (i.e. higher CONCEPT scores). This is consistent with our previous model of the role of social conceptual representations in self-blame (23) and with the finding that the right ATL represented conceptual meaning irrespective of emotional context (46). It is unclear whether lower BOLD activation in the right ATL represents a vulnerability factor for developing MDD or is modulated by mood state, but the association between CONCEPT scores and psychosocial functioning scores, which include symptoms, suggests that it is at least partially modulated by mood state.

Our results implicate the right ATL in depressogenic self-blame-selective conceptual overgeneralization, but the ATL is a large, heterogeneous area containing graded functional specialized subregions (10, 47). Our peak was in anterior BA20, which was previously associated with conceptual representations in general including social concepts (12). However, our cluster extended into BA36, i.e. the perirhinal cortex, which has previously been found to be important for recognizing novel and unique objects (48). Previous work has shown that the perirhinal cortex is involved in conceptual representations of concrete objects irrespective of modality (49). The more inferior right anterior temporal lobe regions may be related to perceptual aspects of person knowledge, including facial and other identifying information, to allow access to semantic information about the unique identity of the individual (13, 50). However, multiple semantic dementia and temporal lobe epilepsy resection studies have found bilateral ATL lesions to be associated with semantic knowledge for faces, potentially with a small right-hemispheric lateralization effect (51–53). We speculate that individuals who more strongly activated right perirhinal semantic representations when blaming themselves, were better able to conceptually differentiate because of better integrating distributed semantic information within the ATL. In other words, self-blame-selective conceptual overgeneralization might be partly driven by a breakdown in the link between abstract social conceptual and familiarity memory systems.

Moreover, we observed a side peak in BA38 related to individual differences in self-blame-selective conceptual overgeneralization, with a more pronounced inter-individually consistent bilateral superior ATL activation across the self- and other-blame conditions. These findings are consistent with suggestions that there is an extended ATL network with conceptual knowledge hubs, such as the “hub-and-spoke” theory of conceptual knowledge (11, 54, 55). This theory posits that the bilateral ATL represents a “hub” that brings together sensory specific and semantic associations which are distributed across the brain (“spokes”). There is evidence of graded functional specialization between and within the ATLs (47, 55). However, more research is required to explain the relative contribution of the different subregions to the representation of conceptual knowledge and their role in MDD pathophysiology.

The subgenual frontal region, shown to be important for MDD pathophysiology (56–58) and directly connected with superior ATL, was hypothesized to play a key role in conceptual-emotional integration when attributing blame (7). Accordingly, we observed that individuals with higher self-blame-selective activation of the right anterior subgenual cingulate (BA24) exhibited more negative CONCEPT x VALENCE scores, suggestive of depressogenic conceptual-emotional integration. Higher subgenual frontal activity for self-blame has been associated with proneness to experiencing feelings of guilt, irrespective of MDD history (18, 32). These findings are in keeping with the anterior subgenual cingulate’s hypothesized role in representing social agency, which is key to attributing blame and thus determines the emotional relevance of appraised actions (23). Individuals with a high interdependence of conceptual overgeneralization and negative emotional valence (i.e. more negative CONCEPT x VALENCE scores) are likely to have attributed more blame when conceptually overgeneralizing, thus explaining more negative valence ratings.

Unexpectedly, we also observed a BOLD effect in the right primary motor area which was negatively associated with conceptual-emotional integration. The activation of the right motor cortex is unlikely to have been related to button press or motion, as only right-handed individuals were included in this study and, therefore, one would have expected left motor cortex activity. We speculate that the motor cortex was activated because of action descriptions activating motor areas, as part of implicit imagery of the action (59). Previous research has shown that motor cortices are activated by verbal stimuli, although it is unclear if this is due to semantic representation or through association (60, 61). It is also plausible that the right primary motor area is relevant in self-agency representation, linked to subjective feelings of motor control (62, 63)

On a cautionary note, this study only probed self-blame-selective conceptual overgeneralization and did not assess other forms of overgeneralization, such as overgeneral autobiographical memory (39) or more contextualized negative self-overgeneralization as measured on the Attitudes Towards Self Scale (64). Previous work has demonstrated that it is likely that different overgeneralization processes contribute to the psychopathology of depression (42, 43). However, it will be important to investigate if and how these processes interact. For example, a recent meta-analysis showed that autobiographical memory overgeneralization persists following remission from MDD (65) and, given our finding of persistent self-blame-selective conceptual overgeneralization, future research should investigate their interaction.

Moreover, despite the clear difference between the rMDD and control groups, there was no association between self-blame-selective conceptual overgeneralization and recurrence risk or number of previous episodes. Further research is required to determine whether self-blame-selective conceptual overgeneralization represents a “scar” effect of a previous depressive episode (66, 67) or a correlate of a primary vulnerability to MDD. This could be probed in people with high vs. low risk of MDD, for instance, based on family history.

## Conclusion

Taken together, the MDD group displayed greater self-blame-selective conceptual overgeneralization and a stronger influence of conceptual overgeneralization on emotional intensity (i.e. more interdependent conceptual-emotional integration), compared to the control group. Self-blame-selective conceptual overgeneralization was associated with lower proneness to externalized anger across groups and poorer psychosocial functioning within the MDD group. In contrast, there was no association with recurrence risk, suggesting conceptual overgeneralization as a primary vulnerability trait marker. Confirming the role of the right ATL in individual differences in conceptual overgeneralization, self-blame-selective conceptual overgeneralization was associated with lower BOLD activation in the right ATL. Conversely, depressogenic conceptual-emotional integration correlated with right anterior subgenual cingulate and right primary motor cortex activations, which is consistent with their previously prosed role in social and motor agency representations, respectively. Future studies are needed to confirm the hypothesis that self-blame-selective conceptual overgeneralization characterizes a distinct neurocognitive subtype of primary MDD vulnerability which may be modulated by depressive state and thus serves as a personalized treatment target.

## Data Availability

All data produced in the present study are available upon reasonable request to the authors.

https://kcl.figshare.com/articles/dataset/Development_of_Cognitive_and_Imaging_Biomarkers_Predicting_Risk_of_Self-Blaming_Bias_and_Recurrence_in_Major_Depression/16473870

## Acknowledgments

This study was funded by an MRC Clinician Scientist Fellowship to RZ (G0902304). RZ and AL were partly funded by the National Institute for Health Research (NIHR) Biomedical Research Centre at South London and Maudsley NHS Foundation Trust and King’s College London. The views expressed are those of the authors and not necessarily those of the NHS, the NIHR or the Department of Health. RZ, CS, and AL were supported by a Medical Research Council grant (ref: MR/T017538/1). JM was supported by the LABS-D’Or Hospital Network, Rio de Janeiro, Brazil. DF was supported by a KCL/IDOR Pioneer Science Fellowship, funded by Scients Institute and the IDOR Pioneer Science Initiative. We thank all the participants for volunteering their time for this study.

## Credit statement

**DF**: Conceptualization, Methodology, Formal analysis, Data curation, Writing - Original Draft, Visualization. **AL**: Software, Formal analysis, Writing - Review & Editing. **CS**: Conceptualization, Methodology, Writing - Review & Editing. **JM**: Conceptualization, Methodology, Writing - Review & Editing. **MLR**: Conceptualization, Methodology, Writing - Review & Editing, Funding acquisition. **RZ**: Conceptualization, Methodology, Formal analysis, Data curation, Writing - Review & Editing, Supervision, Project administration, Funding acquisition.

## Rights retention

For the purposes of open access, the authors have applied a Creative Commons Attribution (CC BY) license to any Accepted Author Manuscript version arising from this submission.

