## Supplementary material for "Self-blame-selective social conceptual overgeneralization and vulnerability to depression": Fennema_etal_MedRxiv_Conceputal_overgeneralization_rMDD_Supplement_v2.docx

Diede Fennema^1^, Andrew J. Lawrence^1^, Catherine A. Spilling^1^, Jorge Moll^2^,

Matthew A. Lambon Ralph^3^ & Roland Zahn^1,2,4*^

*^1^ Centre of Affective Disorders, Institute of Psychiatry, Psychology & Neuroscience, Department of Psychological Medicine, King’s College London, London, UK*

*^2^ Cognitive and Behavioral Neuroscience Unit, IDOR Pioneer Science Initiative, D’Or Institute for Research and Education (IDOR), Rio de Janeiro, Brazil*

*^3^ MRC Cognition and Brain Sciences Unit, University of Cambridge, Cambridge, UK*

*^4^ National Service for Affective Disorders, South London and Maudsley NHS Foundation Trust, London, UK*

* Corresponding author

Professor Roland Zahn (see address above)

Keywords: *social concepts; moral emotions; major depression; self-blame; fMRI*

***Parts of this Supplementary Online Content have been adapted from a previously published one in JAMA Psychiatry (doi: 10.1001/jamapsychiatry.2015.1813).***

### **Supplementary Methods**

#### *Additional inclusion criteria*

The remitted major depressive disorder (rMDD) group had the following additional inclusion criteria: at least one past moderate or severe major depressive episode (MDE; classified according to International Classification of Diseases (1)), and at least one past episode with a minimum duration of two months, where depressive symptoms were not caused by comorbid Axis-I disorders.

Both groups were right-handed, native English speakers, with normal or corrected-to-normal vision.

#### *Additional exclusion criteria*

The following exclusion criteria applied to both groups: history of alcohol or substance abuse, MRI contraindications, psychotropic medication or psychotherapy during the study, significant psychosocial impairment as an indicator of a possible personality disorder (assessed on the Global Assessment of Functioning scale (GAF) (2)), Montgomery-Åsberg Depression Rating Scale (3) (MADRS) score > 10, current self-harming behavior, clinically relevant MRI abnormalities, developmental disorders, learning disabilities, Addenbrooke’s Cognitive Exam-R score < 88 (4) (for participants > 50 years of age), neurological illness or physical illnesses known to significantly alter brain function or blood flow.

The control group had the following additional exclusion criteria: no first-degree family history of MDD, bipolar disorder or schizophrenia.

#### *Recruitment and clinical assessment*

Participants were recruited using online and print advertisements as part of the UK Medical Research Council-funded “Development of Cognitive and Imaging Biomarkers Predicting Risk of Self-Blaming Bias and Recurrence in Major Depression” project (5). As in our previous study (6), eligibility was assessed via phone pre-screening and subsequent in-person assessment using the Structured Clinical Interview-I (SCID-I) for DSM-IV (2), with the decision about final inclusion made by a clinical psychiatrist. All investigators received training and showed excellent inter-rated reliability (5). Participants were followed up clinically at 3, 6 and 14 months either in person or over the phone using the Longitudinal Interval Follow-up Evaluation (7) to determine their Psychiatric Status Rating (PSR), with 1=no symptoms, 2=mild symptoms causing no relevant impairment or distress, 3=mild symptoms causing no more than moderate distress/impairment, 4=major symptoms not meeting full MDE criteria, 5=symptoms meeting full MDE criteria, and 6=most severe forms of MDE. Participants were classified as stable remission (n=34), subthreshold symptoms (n=9) and recurring episode (n=15) based on their highest PSR over the worst two weeks of the follow-up period (stable remission=PSR 1-3 and not requiring treatment, subthreshold symptoms=PSR 3 and requiring treatment or PSR 4, recurring episode=PSR 5-6).

#### *Conceptual Social Knowledge Differentiation task – positive stimuli*

The Conceptual Social Knowledge Differentiation (CSKD) task has two versions, one comprising negative social behavior stimuli and one comprising positive social behavior stimuli. The CSKD version with negative stimuli has been described in detail in the main manuscript. Ninety-three participants (57 rMDD and 36 control) also completed the CSKD task with positive stimuli. In this version of the task, participants were shown descriptions of positive social behavior which differed in their agency (30 self-agency items and 30 other-agency items). For example, in the self-agency condition, participants were asked to imagine “[participant’s name] giving [best friend’s name] a gift whilst shopping”; whereas for the equivalent stimuli in the other-agency condition, participants were asked to imagine “[best friend’s name] giving [participant’s name] a gift whilst shopping”. The CONCEPT, VALENCE and CONCEPT x VALENCE scores were calculated from the positive version of the CSKD in the same manner as for the negative version of the task described in the main manuscript.

#### *Value-related Moral Sentiment Task*

The Value-related Moral Sentiment Task (VMST) probes blame-related emotions and has been described in full elsewhere (5, 8). Participants were shown 180 short written statements describing hypothetical social behaviors, in which either the participant (self-agency) or their best friend (other-agency) acts counter to social and moral values. The same 90 social behaviors were used for self- and other-agency conditions. Participants were asked to select the emotion that best described how they would feel given the unpleasant hypothetical situation, from the following options: guilt, shame, contempt/disgust towards self, contempt/disgust towards friend, indignation/anger towards friend, or no feeling/other feeling.

For the measure “anger towards others”, we calculated the proportion of trials in which indignation/anger towards friend was selected in the other-agency condition.

#### *Behavioral analysis*

Mann-Whitney-*U* and Wilcoxon tests were used to test between-group differences where Kolmogorov-Smirnov tests rejected the assumption of normality at α=0.05. Data were standardized to check for univariate outliers (*z* ≥ 2.5 standard deviations from the mean) and Mahalanobis distance was used to check for bivariate outliers (critical threshold, χ^2^[2]=9.21, at *p*=.01) for the rMDD group and control group separately.

Split-half reliability of the CSKD task was assessed using the Spearman-Brown formula after splitting items in each condition into parallel forms based on the alphabetical order of the stimuli, using a first-second half split approach. A Spearman-Brown (SB) coefficient > .8 is considered “good/excellent”, > .7 is “acceptable”, and < .6 is “poor” (9).

#### *fMRI paradigm*

In the self-agency condition, the participant acted towards their best friend (“[participant’s name] does act dishonestly towards [best friend’s name]”), while in the other-agency condition their best friend acted towards them (“[best friend’s name] does act dishonestly towards [participant’s name]”). The same social concepts were used in both conditions (90 trials per condition, 50% negative social behaviors [e.g. “does act dishonestly”] and 50% negated positive behaviors [e.g. “does not act generously”]). A baseline visual fixation of pattern condition (number of trials = 90) was interspersed. Participants were asked to provide the name of their best friend, with whom they were not related and not romantically involved.

#### *MRI acquisition*

All images were acquired on a Phillips 3-Tesla Achieva MRI scanner equipped with an eight-channel head coil. T2*-weighted images were acquired with a sensitivity-encoded echo-planar sequence (repetition time (TR)=2000ms, echo time (TE)=20.5ms, sensitivity encoding factor=2) in three runs of 405 volumes, including five dummy volumes (run time: 13 min 40 sec). Depending on head size, 35 to 40 slices with 3 mm slice thickness were acquired in a continuous ascending order, parallel to the anterior to posterior commissural line, providing images with an 80 x 80 acquisition matrix and reconstructed voxel size 2.29 x 2.29 x 3mm, covering a field of view (FOV) of 220 x 220 x 120mm.

In addition, three-dimensional T1-weighted magnetization-prepared rapid acquisition gradient echo structural images were obtained (160 axial slices, 0.9mm slice thickness; TR=8.4ms, TE=3.9ms, FOV=240 x 191 x 144mm, acquisition matrix=256 x 163 voxels, reconstructed voxel size=0.94 x 0.94 x 0.9mm; flip angle 8 degrees).

#### *fMRI analysis*

After the scanning session, participants rated the degree of unpleasantness associated with each stimulus on a 7-point Likert scale (ranging from 1=not unpleasant, 7=extremely unpleasant). Self- and other-blaming emotion trials for each individual were defined as those trials with unpleasantness ratings at or above that individual’s median rating in the self- and other-agency conditions.

The original researchers made standard pre-processed functional images available, i.e. functional images which were realigned, unwarped and coregistered to the participant’s T1-weighted images, spatially normalized and smoothed with a 6mm Full-Width at Half-Maximum smoothing kernel. At the individual level, blood-oxygen level-dependent (BOLD) effects were modelled for self- and other-agency emotion trials, including only trials for each individual with unpleasantness ratings at or above that individual’s median rating in the self- and other-agency conditions. We modeled null events and six-degree movement parameters as nuisance regressors but chose not to model time and dispersion derivates at the individual level, as previous work had shown that this reduced internal consistency (10).

As described in the main manuscript, we conducted one-sample *t*-tests comparing BOLD and PPI contrast maps for self- vs other-blaming emotions, while modeling CONCEPT, VALENCE and CONCEPT x VALENCE scores as covariates, so that the observed activation is more likely to relate to conceptual-emotional integration as opposed to emotional appraisal or conceptual differentiation alone. In contrast with Green *et al.* (11), we did not include group as a covariate given that group is associated with our covariate of interest, rendering the results unreliable (12).

#### *fMRI ROIs*

The following ROIs were selected, based on previous independent studies showing relevance for social conceptual-emotional integration (see Figure 1).

1. Right hemispheric part of a previously described bilateral *a priori* ATL region, see supplement of Zahn *et al.* (13), created using Brodmann area (BA) 38, 22 and 21 maps from the WFU PickAtlas (14) and restricting them to MNI y-coordinates ≥ -10.
2. *A priori* bilateral DLPFC ROI, based on the Automated Anatomical Labelling atlas (15), described in the supplement of Zahn *et al.* (13). A broader DLPFC ROI was used than the one identified in Green *et al.* (6) to account for individual variability.
3. *A priori* bilateral subgenual anterior cingulate (BA24), as described in Green *et al.* (6), 6mm spheres centered on MNI coordinates: x=+/-4, y=23, z=-5.

### **Supplementary Results**

#### *Split-half reliability*

Split-half reliability of the CONCEPT measure was excellent across the different conditions (negative self-agency: SB=.91; negative other-agency: SB=.92; positive self-agency: SB=.93; positive other-agency: SB=.92). However, split-half reliability of the VALENCE measure was only acceptable in the negative condition (negative self-agency: SB=.73; negative other-agency: SB=.67), and poor in the positive condition (positive self-agency: SB=-.07; positive other-agency: SB=-.17).

#### *Association between negative version CSKD task and clinical measures*

Within the rMDD group, there was a negative association between CONCEPT scores and GAF (*r*[60]=-0.30, *p*=.02; Supplementary Table 1) and a positive association between VALENCE scores and GAF (*r*[60]=.27, *p*=.04; Supplementary Table 1), i.e. those with self-blame-selective conceptual overgeneralization or negative emotional valence biases had lower psychosocial functioning scores. Moreover, CONCEPT x VALENCE scores were positively associated with GAF (*r*[60]=.27, *p*=.04) and negatively associated with Beck Depression Inventory scores (*r*[60]=-.28, *p*=.03), i.e. those with more negative interdependence scores had lower psychosocial functioning scores and higher depressive symptom levels (Supplementary Table 1).

#### *Negative version CSKD task with outliers removed*

When excluding seven uni- and bivariate outliers (rMDD: 4; control: 3), the rMDD group continued to display self-blame-selective overgeneralization, i.e. selecting more concepts in the negative self-agency condition (M=1.61, SD=0.60) than in the other-agency condition (M=1.50, SD=0.54; *t*[55]=3.13, *p*=.003), while the control group selected a similar number of concepts in the self-agency condition (M=1.41, SD=0.46) as in the other-agency condition (M=1.41, SD=0.52; *t*[32]=0.00, *p=*1.00).

Furthermore, when excluding the outliers, group differences remained as before outlier exclusion on CONCEPT (rMDD, M=0.11, SD=0.27; control, M=0.00, SD=0.18; *t*[85.9]=2.37, *p*=.02) and CONCEPT x VALENCE scores (rMDD, M=-0.08, SD=0.18; control, M=-0.02, SD=0.06; *t*[74.6]=-2.31, *p*=.02). As in our primary analysis, there were no group differences in VALENCE scores (rMDD, M=-0.14, SD=0.42; control, M=-0.04, SD=0.29; *t*[85.0]=-1.30, *p*=.20).

#### *Positive CSKD task version results*

For the positive CSKD version, on average both groups selected similar numbers of concepts in the self-agency condition (rMDD: M=1.69, SD=0.70, control: M=1.53, SD=0.69) as in the other-agency condition (rMDD: M=1.72, SD=0.73; *t*[56]=-1.18, *p*=.25; control: M=1.56, SD=0.71; *t*[35]=-1.17, *p*=.25). Removal of nine uni- and bivariate outliers (rMDD: 5; control: 4) did not alter these findings. The rMDD group still selected on average similar numbers of concepts in the self-agency condition (M=1.66, SD=0.72) compared to the other-agency condition (M=1.68, SD=0.74; *t*[51]=-1.07, *p*=.29), as did the control group (self-agency: M=1.44, SD=0.59; other-agency: M=1.45, SD=0.61; *t*[31]=-1.16, *p*=.26).

There were no significant group differences for either CONCEPT (rMDD: M=-0.03, SD=0.22; control: M=-0.02, SD=0.11; *t*[86.7]=-0.38, *p*=.70), VALENCE (rMDD: M=0.06, SD=0.33; control: M=0.04, SD=0.35; *t*[91]=0.29, *p*=.76) or CONCEPT x VALENCE scores (rMDD: M=0.03, SD=0.11; control: M=0.002, SD=0.02; *t*[62.0]=1.56, *p*=.12) for the positive version of the CSKD task. With outlier exclusion, there were no significant group differences for either CONCEPT (rMDD: M=-0.02, SD=0.15; control: M=-0.01, SD=0.07; *t*[76.4]=-0.35, *p*=0.73) or VALENCE (rMDD: M=0.05, SD=0.27; *t*[82]=0.77, *p*=0.45). However, there was a significant group difference for CONCEPT x VALENCE (rMDD: M=0.01, SD=0.05; control: M=-0.001, SD=0.01; *t*[54.9]=2.03, *p*=.05).

### **Supplementary Tables**

**Table S1 | Subclusters anterior temporal lobe BOLD activation related to conceptual overgeneralization (n = 75).**

|  |  |  |  | *MNI peak coordinates* | | |  |  |
| --- | --- | --- | --- | --- | --- | --- | --- | --- |
| Hemi-sphere | Region | Cluster size (voxels) | Brodmann Area | x | y | z | *t* statistic | Uncorrected *p* value |
| BOLD negative effect of CONCEPT scores: | | | | | | | | |
| right | Anterior temporal lobe | 39 | 20/36 | 40 | -2 | -34 | 4.58 | .000^a^ |
|  |  |  | 38 | 46 | 16 | -18 | 3.28 | .001 |
| ^a^ Region surviving voxel-based FWE correction over *a priori* right anterior temporal lobe region-of-interest (*p* = .006) as described in the supplement of Zahn *et al.* (13).  CONCEPT is calculated as the difference in mean number of concepts chosen in the self- minus the other-agency as an index of conceptual overgeneralization. BOLD = blood-oxygen level-dependent; FWE = Family-Wise Error; MNI = Montreal Neurological Institute. | | | | | | | | |

**Table S2 | Association between CSKD measures and clinical characteristics remitted MDD.**

|  |  | CONCEPT | VALENCE | CONCEPT x VALENCE |
| --- | --- | --- | --- | --- |
| GAF | *r* | -.30* | .27* | .27* |
|  | *p*-value | .02 | .04 | .04 |
| BDI | *r* | .16 | -.01 | -.28* |
|  | *p*-value | .22 | .92 | .03 |
| MADRS | *r* | .20 | -.17 | -.19 |
|  | *p*-value | .12 | .20 | .15 |
| Number of lifetime MDEs | *r* | .07 | .12 | -.05 |
|  | *p*-value | .60 | .35 | .69 |
| * significant at *p* < .05 threshold, two-tailed.  Based on n=60 participants. CONCEPT is calculated as the difference in mean number of concepts chosen in the self- minus the other-agency as an index of conceptual overgeneralization; VALENCE is calculated as the difference in mean (un)pleasantness ratings in the self- minus the other-agency as an index of emotional valence bias; and CONCEPT x VALENCE is the multiplication of CONCEPT and VALENCE scores. CSKD = Conceptual Social Knowledge Differentiation; MDD = Major Depressive Disorder; GAF = Global Assessment of Functioning scale; BDI = Beck Inventory Scale of Depression; MADRS = Montgomery-Åsberg Depression Rating Scale; MDE = Major Depressive Episode. | | | | |

**Table S3 | Baseline characteristics by group self-blame-selective conceptual overgeneralizer vs. non-self-blame-selective conceptual overgeneralizer.**

|  | Self-blame-selective conceptual overgeneralizer | Self-blame-selective non-conceptual overgeneralizer | Comparison |
| --- | --- | --- | --- |
|  | n = 40 | n = 56 |  |
| Age | 36.4 ± 14.4; 18 - 64 | 35.7 ± 13.3; 20 - 62 | *t*(94) = .24, *p* = .81 |
| Gender |  |  | χ^2^ (1, 96) = 1.88, *p* = .17 |
| Female | 29 (73%) | 33 (59%) |  |
| Male | 11 (28%) | 23 (41%) |  |
| Education (years) | 16.9 ± 2.3; 12 -22 | 17.3 ± 2.3; 12 - 25 | *t*(94) = -0.71, *p* = .48 |
| MADRS | 1.0 ± 1.4; 0 - 4 | 0.8 ± 1.4; 0 - 4 | *U*(96) = 1010.0, *p* = .32 |
| BDI | 3.1 ± 3.9; 0 - 15 | 2.7 ± 3.3; 0 - 12 | *U*(96) = 1046.0, *p* = .57 |
| GAF | 85.8 ± 6.1; 70 - 90 | 87.5 ± 5.0; 70 - 91 | *U*(96) = 1300.0, *p* = .11 |
| Classification |  |  | χ^2^ (1, 96) = 2.93, *p* = .09 |
| MDD | 29 (73%) | 31 (55%) |  |
| Control | 11 (28%) | 25 (45%) |  |
| Based on n=96 participants. “Self-blame-selective conceptual overgeneralizer” is defined as participants with positive CONCEPT scores and “non-self-blame-selective conceptual overgeneralizer” is defined as participants with CONCEPT scores equal to or less than 0. CONCEPT is calculated as the difference in mean number of concepts chosen in the self- minus the other-agency as an index of conceptual overgeneralization.  Means, standard deviations and range are reported (*M ± SD; minimum – maximum).* Percentages may not add up to 100 due to rounding. MADRS = Montgomery-Åsberg Depression Rating Scale; BDI = Beck Depression Inventory; GAF = Global Assessment of Functioning; MDD = Major Depressive Disorder. | | | |

**Supplementary Figure Legends**

**Figure S1 | Visualization of anterior temporal lobe BOLD cluster related to conceptual overgeneralization.**

The figure shows a cropped section through the bilateral anterior temporal lobe clusters, displayed using MRIcron (16) at a Family-Wise Error corrected voxel-level threshold of *p* = .05, with no cluster-size threshold. The color bar represents *t* values and the x denotes the MNI x-coordinate of the slice. The activation represents the consistent BOLD effects in the anterior temporal lobes across the self- and other-agency conditions. BOLD = blood-oxygen level-dependent; MNI = Montreal Neurological Institute.
