## Supplementary figures and images for "Self-blame-selective social conceptual overgeneralization and vulnerability to depression"

### FigureS1.tiff

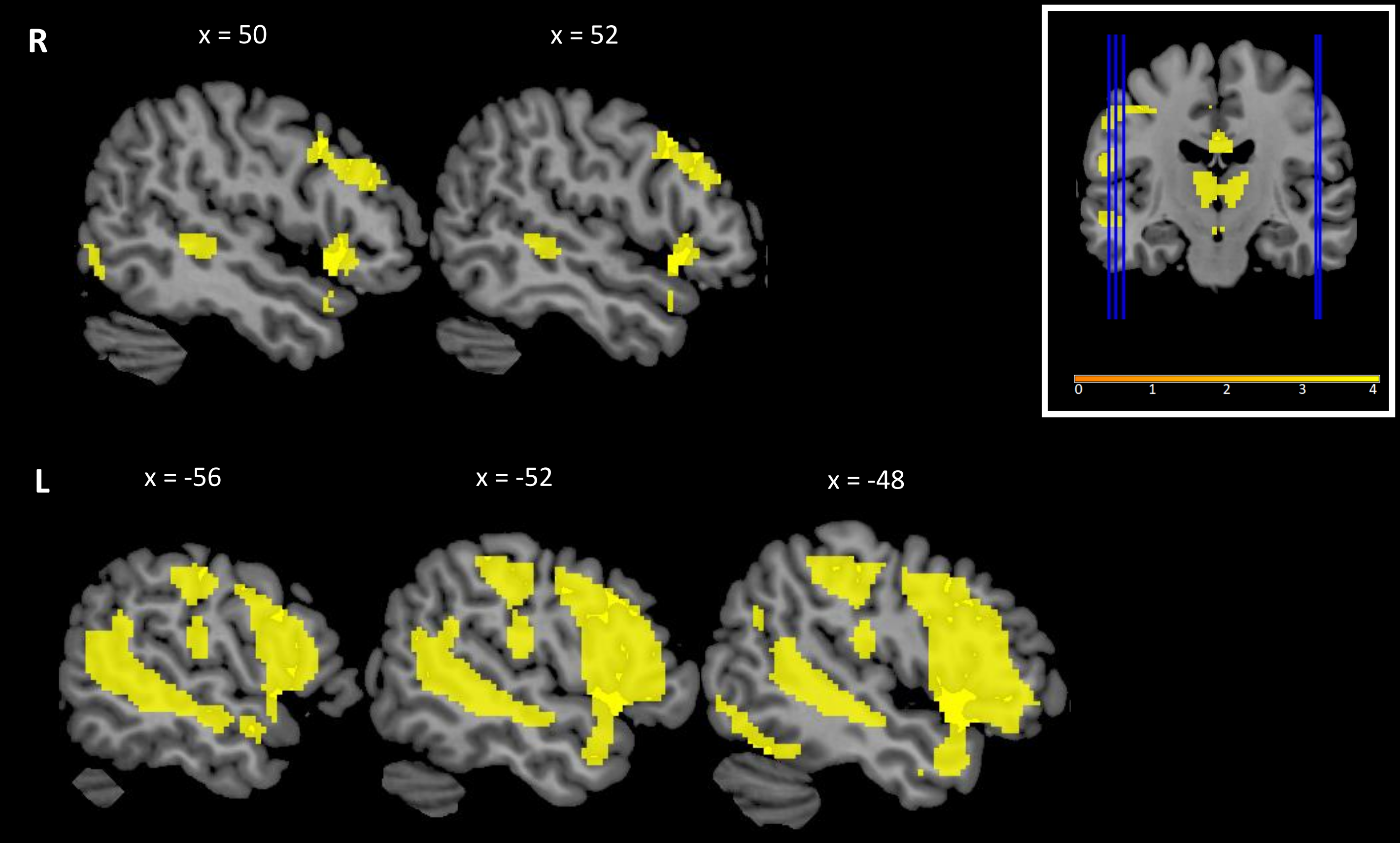
